# Development of a symptom-based severity score anchored to health-related quality of life post-COVID-19 within the population-based EPILOC cohorts

**DOI:** 10.64898/2026.06.08.26355135

**Authors:** Raphael S. Peter, Lisamaria Sedelmaier, Alexandra Nieters, Claudia Schilling, Lynn Matits, Siri Göpel, Uta Merle, Jürgen M. Steinacker, Dietrich Rothenbacher, Winfried V. Kern, the EPILOC Omicron Study Group

## Abstract

**Purpose:** Because simple symptom counts treat all symptoms as equally important and may not adequately capture the HRQoL impact of heterogeneous post-COVID-19 symptoms, we aimed to develop an HRQoL-anchored symptom severity score providing an interpretable measure of post-COVID-19 disease burden.

**Methods:** Baseline data from the population-based EPILOC and EPILOC Omicron surveys (adults aged 18–65 years) were used to develop a symptom-based severity score anchored to physical and mental HRQoL assessed with the SF-12. A two-stage modelling approach was applied to identify HRQoL-relevant symptoms and to derive symptom-specific weights for physical and mental component scores, incorporating 30 ordinal symptom severity variables. Symptom-specific weights were extracted to compute physical, mental, and composite severity scores. Score interpretation was examined using external reference measures, including EPILOC case status, self-reported health recovery, and functional consequences.

**Results:** A total of 19,004 participants (mean age 44.3 years, 59.6% female) were included. Sixteen symptoms contributed to the physical and eleven to the mental HRQoL score, with a limited subset accounting for most of the HRQoL loss. Severity scores were heavily right-skewed, with 50.6% of participants showing no measurable HRQoL impairment. Higher scores correlated with lower self-reported recovery, and increased probability of rehabilitation use and health-related changes in working time, supporting convergent and criterion-related validity.

**Conclusions:** This study introduces a transparent, HRQoL-anchored symptom severity score that measures graded post-COVID-19 burden beyond simple symptom counts. The score may be particularly suited for longitudinal assessment of recovery trajectories.

**Plain English summary:** Many people report ongoing symptoms after a COVID-19 infection, but it is hard to explain how much these symptoms impact daily life and well-being. Simply counting symptoms is not effective because some are more severe or disruptive than others.

This study focuses on how to measure the overall effect of post-COVID symptoms on people’s quality of life in a clear and meaningful way. The goal was to develop a score that shows how strongly different symptoms are associated with limitations in physical and mental health.

Using data from over 19,000 participants, we connected symptom severity to well-established measures of physical and mental quality of life. We found that only a few symptoms had strongly impaired daily functioning, while many others had little or no measurable effect. The resulting score clearly differentiated individuals who had fully recovered from those with ongoing health issues.

Overall, this study demonstrates that a quality-of-life-based symptom score offers a clearer and more useful view of post-COVID health compared to simple symptom counts. Such a score can help researchers and healthcare providers better understand recovery and ongoing health needs after COVID-19.

## Introduction

Post-COVID-19 sequelae are increasingly recognised as a relevant public health concern, with a substantial proportion of individuals experiencing persistent or new symptoms weeks or months after initial SARS-CoV-2 infection [1]. These symptoms can affect multiple organs and vary greatly in type, severity, and duration [2,3]. Beyond symptom presence, impairment in daily functioning and quality of life has become a key outcome in post-COVID-19 research and a central concern for patients, healthcare providers, and health systems. In a recent post-COVID-19 umbrella review, 55% of all systematic reviews analysed included a measure of health-related quality of life (HRQoL) [4].

This growing emphasis on HRQoL highlights the need for appropriate ways to summarise symptom burden. A common approach in both clinical and epidemiological research is to summarise symptom burden by simple counts of reported symptoms [5–9]. While practical, such unweighted symptom counts have significant limitations from a measurement perspective: they assume all symptoms are equally important, overlook differences in individual symptom severity, and are only loosely linked to functional health outcomes.

In the context of patient-reported outcome research, symptom-based severity measures should be anchored to validated HRQoL instruments to ensure clarity, patient and clinical relevance [10,11]. Instead of establishing a new post-COVID disease entity or case definition, the goal is to operationalise symptom burden in a way that accurately reflects its observed impact on functional health.

We therefore aimed to develop a symptom-based severity score for post-COVID-19 syndrome that is explicitly anchored to HRQoL. The score combines information on both symptom presence and graded symptom severity, allows differential weighting of symptoms based on their empirical association with HRQoL, provides an interpretable measure of individual burden that will be suitable for longitudinal assessment of recovery trajectories, and distinguishes symptoms with a meaningful impact on HRQoL from those with limited functional significance.

## Methods

### Study design and data sources

EPILOC (Epidemiology of Long Covid) is a population-based longitudinal observational study conducted in the federal state of Baden-Württemberg, Germany. The baseline assessment included adults aged 18 to 65 years with a PCR confirmed, reported SARS-CoV-2 infection occurring between 1 October 2020 and 1 April 2021, notified to public health offices. Participants were invited six to twelve months after the index infection via local public health authorities and completed a standardised questionnaire following written informed consent. Details of the EPILOC baseline assessment have been reported previously [12].

EPILOC Omicron recruited individuals from the same source population who were infected between 15 June and 15 July 2022, using otherwise identical inclusion criteria and study procedures. At the time of recruitment, SARS-CoV-2 infections in Germany were still subject to mandatory notification to local public health authorities, although not all infections could be PCR-confirmed due to limited testing capacity [13].

Both surveys used identical questionnaires, collecting information on sociodemographic characteristics, SARS-CoV-2 reinfections, vaccination status, general health, working capacity, and health-related quality of life (HRQoL), assessed using the Short Form Health Survey (SF-12). In addition, participants reported the presence of 30 predefined symptoms before the index infection, during the acute phase of their SARS-CoV-2 infection, and at the time of questionnaire completion. Severity of persistent or newly occurring symptoms was assessed using an ordinal Likert-type scale (none, light, moderate, severe). In EPILOC Omicron only, post-exertional malaise (PEM) was assessed as fatigue beyond 14 h after physical or mental exertion yes/no. A detailed description of symptom prevalences in the EPILOC cohorts has been reported previously and is therefore not shown again [12,13].

### Score construction

To derive a symptom-based severity score for post-COVID-19 sequelae, we applied a two-stage modelling approach separately for mental and physical HRQoL [14]. In the first stage, an additive model was fitted using all 30 ordinal symptom severity variables as linear terms, along with covariates including age (modelled as a spline), sex, education, smoking status, and survey (Omicron vs. wild-type). Symptoms were retained for further modelling if they showed a statistically significant (p < 0.05) and clinically relevant association with HRQoL, operationalised as a minimum decrease of 0.1 points per severity level.

In the second stage, selected symptoms were refitted using monotonically decreasing smooth functions [15] to ensure measurement plausibility, such that increasing symptom severity could not result in improved HRQoL. Physical and mental SF-12 component scores were modelled separately to maintain the conceptual distinction between physical and mental dimensions of HRQoL. The estimated partial effects were extracted to obtain symptom-specific weights. These represent estimated reductions in SF-12 component scores associated with increasing symptom severity.

To compute individual symptom severity scores, only symptoms that were not present prior to infection were considered. Limiting the score to symptoms that appeared after infection was intended to approximate symptom burden attributable to post-acute sequelae rather than overall morbidity. The symptom weights – representing estimated reductions in HRQoL – were used in positive (absolute) form so that the resulting scores reflect positive symptom burdens. The final composite severity score was calculated as the Euclidean distance between the physical and mental symptom scores:

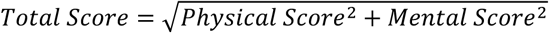

This composite score provides an overall estimate of HRQoL loss attributable to post-acute symptoms, while still allowing separate analysis of physical and mental scores.

### Validation against external reference measures

To aid interpretation and validate the results, the derived severity scores were examined in relation to several external reference measures. Score distributions were compared across EPILOC post-COVID-19 syndrome (PCS) case status categories (PCS cases, fully recovered controls, and participants with some persisting sequalae not sufficient to meet the PCS case definition ‘intermediate’) [16] to determine whether the score distinguishes between groups of different symptom burden.

Second, convergent validity was evaluated by analysing the association between severity scores and self-reported percentage of health recovered since SARS-CoV-2 infection. Spearman’s rank correlation coefficient was used to measure the strength of the association between physical, mental, and total scores and perceived recovery.

Third, criterion-related validity was assessed by examining the relations between severity scores and indicators of functional outcomes, such as planned or attended rehabilitation or health-related changes in working time. These associations were modelled using logistic regression with spline functions without monotonicity constraints to estimate the probability of functional outcomes across different score levels, with results shown as predicted probabilities and 95% confidence intervals. R version 4.5.0 was used for the statistical analyses [17].

## Results

A total of 19,004 participants with complete information on SF-12, all 30 symptoms, and relevant covariates were used for the model-based score construction (**Table 1**). These data provided the basis for estimating symptom-specific contributions to physical and mental HRQoL.

**Table 1.** Characteristics of the study population.

|  | **Overall**<br>N = 19,004 | **EPILOC**<br>N = 8,540 | **EPILOC Omicron**<br>N = 10,464 |
| --- | --- | --- | --- |
| Age (years), mean (sd) | 44.3 (13.5) | 43.1 (13.7) | 45.2 (13.2) |
| Female sex, n (%) | 11,326 (59.6) | 4,902 (57.4) | 6,424 (61.4) |
| General school-leaving qualification, n (%) |  |  |  |
| None | 107 (0.6) | 72 (0.8) | 35 (0.3) |
| Basic | 1,797 (9.5) | 1,007 (11.8) | 790 (7.5) |
| Intermediate secondary | 5,424 (28.5) | 2,736 (32.0) | 2,688 (25.7) |
| University entrance qualification | 11,676 (61.4) | 4,725 (55.3) | 6,951 (66.4) |
| Smoking status, n (%) |  |  |  |
| Never | 12,883 (67.8) | 5,629 (65.9) | 7,254 (69.3) |
| Former | 4,270 (22.5) | 2,051 (24.0) | 2,219 (21.2) |
| Current | 1,851 (9.7) | 860 (10.1) | 991 (9.5) |
| SF-12 component scores, mean (sd) |  |  |  |
| Physical | 50.7 (8.1) | 50.1 (8.5) | 51.2 (7.8) |
| Mental | 49.3 (10.1) | 48.9 (10.3) | 49.6 (9.9) |
| EPILOC PCS case definition, n (%) |  |  |  |
| Control | 9,874 (53.3%) | 3,527 (41.9%) | 6,347 (62.8%) |
| Intermediate | 5,046 (27.2%) | 2,624 (31.2%) | 2,422 (24.0%) |
| Case | 3,606 (19.5%) | 2,264 (26.9%) | 1,342 (13.3%) |
| Unknown | 478 | 125 | 353 |
| Self-reported % health recovered, n (%) |  |  |  |
| 100 | 11,163 (60.3%) | 4,017 (47.7%) | 7,146 (70.7%) |
| 90 | 3,613 (19.5%) | 2,079 (24.7%) | 1,534 (15.2%) |
| 80 | 2,080 (11.2%) | 1,277 (15.2%) | 803 (7.9%) |
| 70 | 871 (4.7%) | 538 (6.4%) | 333 (3.3%) |
| 60 | 346 (1.9%) | 229 (2.7%) | 117 (1.2%) |
| 50 | 208 (1.1%) | 136 (1.6%) | 72 (0.7%) |
| 40 | 90 (0.5%) | 55 (0.7%) | 35 (0.3%) |
| 30 | 84 (0.5%) | 45 (0.5%) | 39 (0.4%) |
| 20 | 37 (0.2%) | 20 (0.2%) | 17 (0.2%) |
| 10 | 12 (0.1%) | 9 (0.1%) | 3 (0.0%) |
| 0 | 17 (0.1%) | 9 (0.1%) | 8 (0.1%) |
| Unknown | 483 | 126 | 357 |
| Functional health consequences, n (%) |  |  |  |
| Planned or attended rehab | 648 (3.4) | 473 (5.6%) | 175 (1.7%) |
| Change in working time due to health reasons<sup>1</sup> | – | 299 (4.0%) | – |

<sup>1</sup> Only assessed in EPILOC

### Symptom weight structure

Of the 30 symptoms evaluated, 16 showed a statistically significant and clinically meaningful negative association with the physical SF-12 component (**Supplemental Table 1**), while 11 symptoms were associated with the mental component (**Supplemental Table 2**). The resulting weight structure indicated that a subset of symptoms contributed disproportionately to HRQoL loss. Rapid physical exhaustion, arthralgia, and shortness of breath had the highest weights in the physical domain, whereas depressive mood showed the strongest contribution to the mental domain. Several complaints, including rapid physical exhaustion, headache, chronic fatigue, concentration difficulties, and altered taste, contributed to both domains, emphasising their broad functional impact.

### Distribution and interpretability of severity scores

The derived severity scores were heavily right-skewed, with 50.6% of participants having a score of 0.0 in both the physical and mental domains, indicating no measurable symptom-related loss of HRQoL. This pattern reflects significant recovery in most individuals, while clearly highlighting a smaller subgroup with ongoing impairment of varying degrees. The joint distribution of physical and mental scores is shown in **Figure 1**.

**Figure 1.**
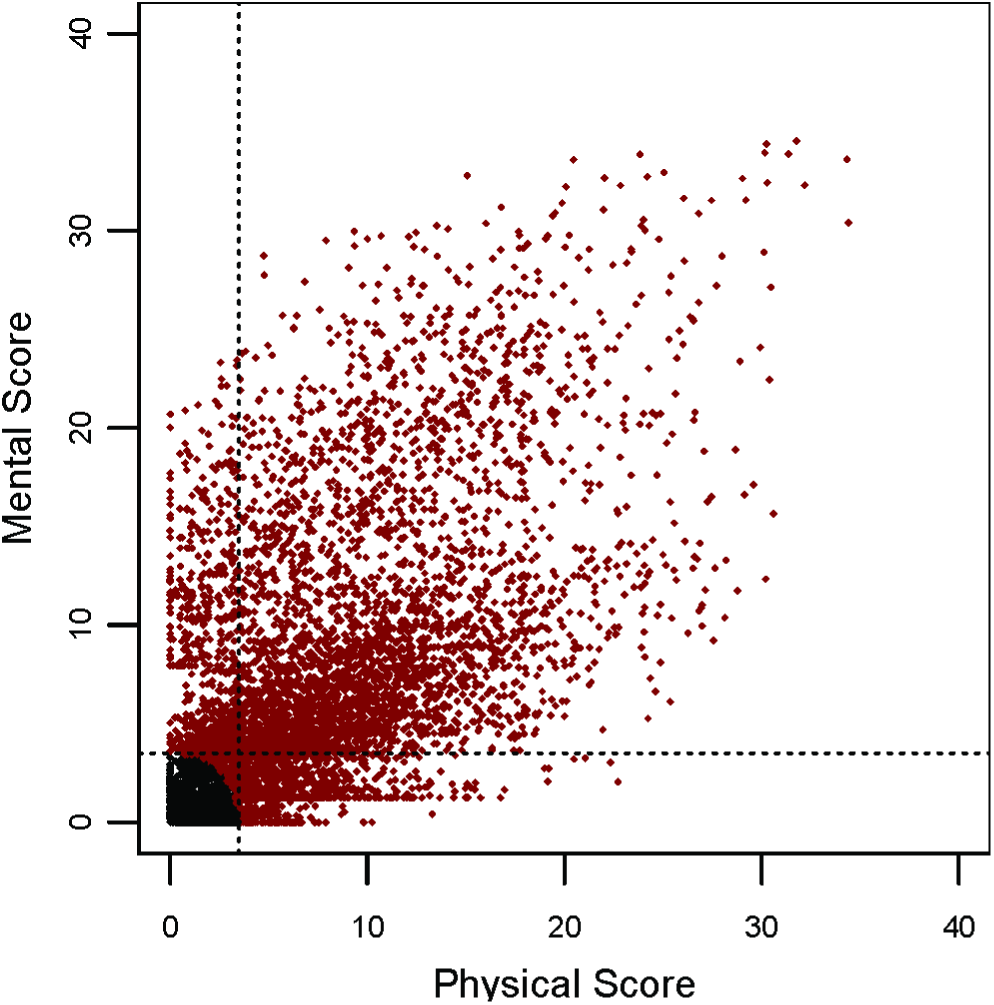
Scatter plot of mental by physical symptom severity scores. Dotted lines represent a cut-off of 3.5 on either scale. Black dots (69%) are <3.5 on the total scale, red dots ≥3.5.

Based on published estimates of minimal clinically important differences for the SF-12, a value of 3.5 points was used as an anchor-based reference for clinically meaningful HRQoL loss in the physical, mental, and total scores. Using this reference, 69% of participants fell below the threshold in all score dimensions. Scores at or above this level were less common among participants from the EPILOC Omicron survey than the EPILOC (wild-type) survey (**Table 2**).

**Table 2.** Prevalence of symptom severity scores ≥3.5 for participants of the Wild-type and Omicron survey.

|  | **EPILOC**<br>N = 8,540 | **EPILOC Omicron**<br>N = 10,464 |
| --- | --- | --- |
| Physical Score $\geq 3.5$, n (%) | 2,739 (32.1) | 1,908 (18.2) |
| Mental Score $\geq 3.5$, n (%) | 2,497 (29.2) | 1,790 (17.1) |
| Total Score $\geq 3.5$, n (%) | 3,319 (38.9) | 2,514 (24.0) |

### Association with reference measures

Score values varied systematically across EPILOC case status categories and levels of self-reported health recovery (**Figure 2**). Median total scores were highest among EPILOC cases, intermediate among participants classified as “intermediate”, and lowest among recovered participants (controls), supporting the discriminative capacity of the score across clinically meaningful groups.

**Figure 2.**
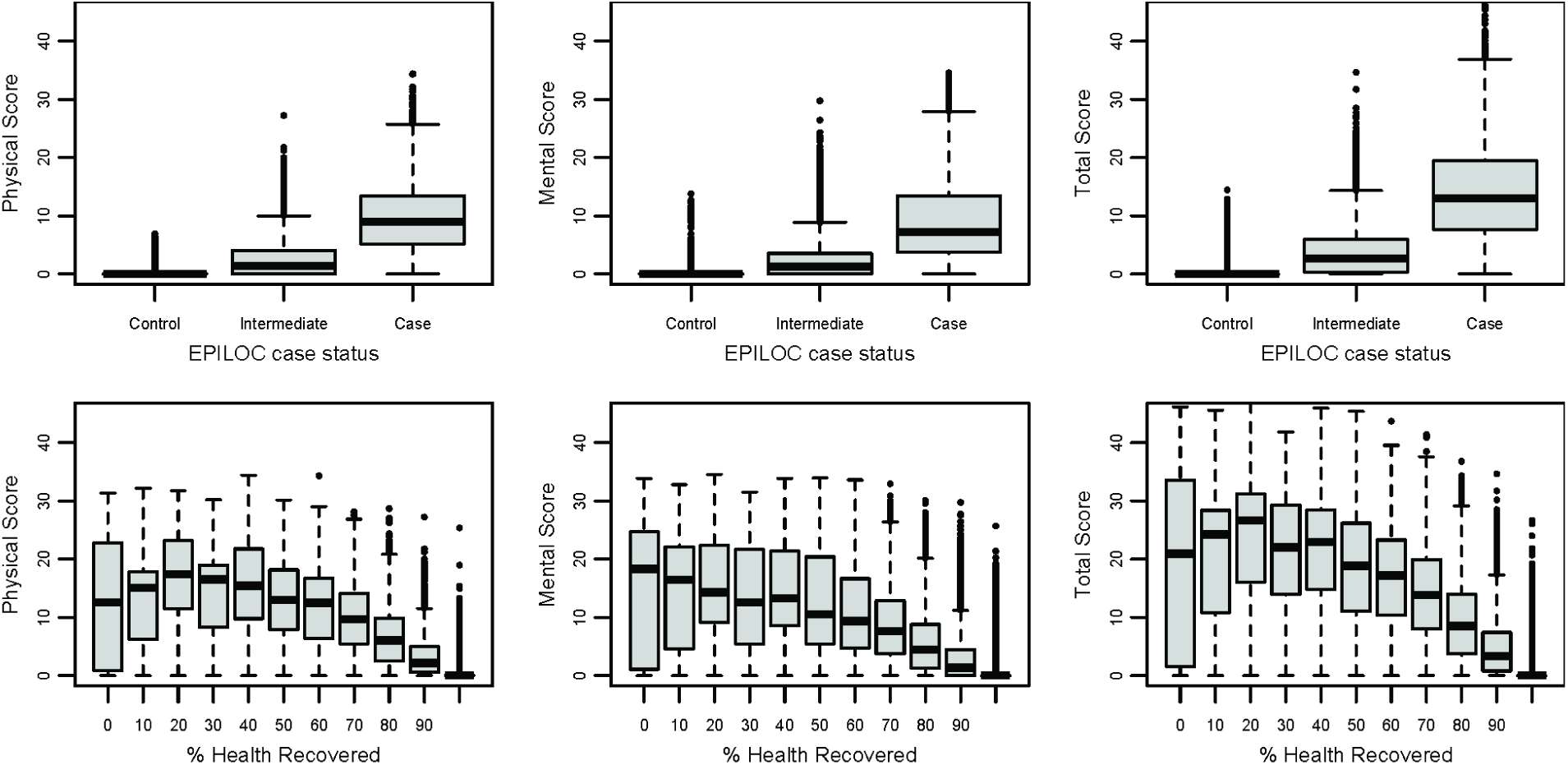
Distribution of symptom severity scores by EPILOC post-covid-19 syndrome case status and self-reported percentage of health recovered.

Convergent validity was demonstrated through strong correlations between severity scores and self-reported percentage of health recovered. The correlations were highest for the physical and total scores (Spearman ρ = −0.75 and −0.73, respectively), and slightly lower for the mental score (ρ = −0.69).

Criterion-related validity was further supported by a dose-response association between increasing score values and the probability of functional consequences, including rehabilitation use and health-related changes in working time (**Figure 3**). These associations were consistently stronger for the physical and total scores than for the mental score alone, indicating a close link between physical symptom burden and observable functional limitations.

**Figure 3.**
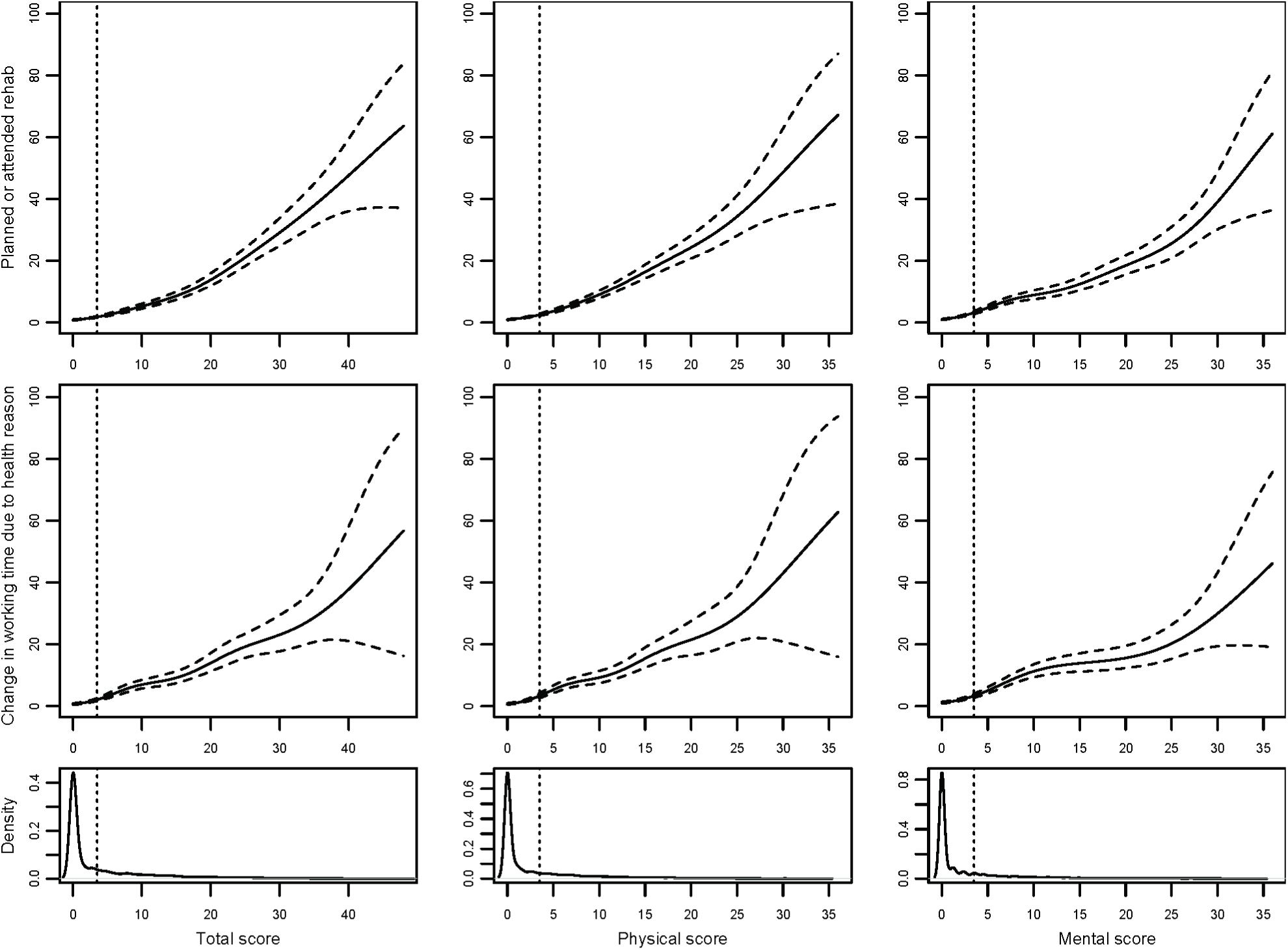
Probability (95%-CI) of functional consequences by the symptom severity scores. The dotted lines represent a 3.5 points cut-off.

## Discussion

In this large population-based study of more than 19,000 adults with prior SARS-CoV-2 infection, we developed a symptom-based severity score anchored to physical and mental HRQoL to quantify symptom burden in post-COVID-19. This work demonstrates how HRQoL-based weighting principles can be transparently operationalised to construct an interpretable severity measure that goes beyond unweighted symptom counts. A key finding is that only a subset of the reported 30 symptoms significantly contributed to reduced HRQol. This empirically derived weighting structure highlights a major limitation of simple symptom counts, which implicitly assume all symptoms hold equal relevance. Clinically, functional impairment may sometimes be driven by a limited number of particularly burdensome symptoms rather than the overall number of reported symptoms, supporting the rationale for weighted HRQoL-based approaches. The symptoms with the greatest contributions – rapid physical exhaustion, arthralgia, and shortness of breath for physical HRQoL, and depressive mood for mental HRQoL – align with previous evidence on post-COVID-19 sequelae [18,19], supporting the construct validity of the score.

Notably, post-exertional malaise (PEM), which is increasingly recognised as a central feature of post-COVID-19 syndrome [20], was not explicitly assessed in the EPILOC baseline assessment. However, several highly weighted symptoms, particularly rapid physical exhaustion and fatigue-related complaints, likely capture aspects of PEM-related burden indirectly. In an exploratory analysis of the EPILCO Omicron subsample, including binary PEM status, the addition of PEM provided only a modest incremental improvement in predicting SF-12 outcomes, suggesting that PEM-related functional impairment may already be partially reflected in the existing symptom structure. Consistent with this interpretation, PCS cases reporting PEM showed higher severity scores than those without PEM (**Supplemental Figure 1**).

Modelling physical and mental HRQoL separately proved informative, as it revealed both domain-specific and cross-domain symptom effects. Symptoms such as chronic fatigue, headache, concentration difficulties, and altered taste affected both domains, reflecting their broad functional relevance. The composite total score, constructed as an integrated summary of physical and mental symptom burden, offers a practical measure of overall HRQoL loss while maintaining the conceptual distinction between health dimensions. Importantly, the availability of domain-specific scores enables flexible application depending on the research or clinical context.

### Distribution, interpretability, and validity of the severity score

The pronounced right skewness of the score distribution, with more than half of participants showing no measurable HRQoL loss, reflects substantial recovery in most individuals after SARS-CoV-2 infection. At the same time, the score clearly identifies a smaller subgroup with persistent and clinically significant impairment. Using established estimates of minimal clinically important differences for the SF-12, a value of 3.5 points served as an anchor-based reference for clinically relevant HRQoL loss [21]. This reference should be viewed as an aid to score interpretation rather than a diagnostic threshold. Comparison with the existing EPILOC case definition demonstrates how the severity score captures gradations of HRQoL impact beyond a simple binary classification.

Several findings support the validity of the proposed measure. Strong correlations with self-reported percentage health recovery provide evidence of convergent validity, while dose-response associations with rehabilitation use and health-related changes in working time support criterion-related validity. The consistently stronger associations observed for the physical and total scores, compared with the mental score alone, emphasise the prominent role of physical symptom burden in driving observable functional consequences.

The lower prevalence of higher severity scores in the EPILOC Omicron cohort compared to the wild-type cohort indicates a reduced average HRQoL impact following Omicron infections. This might be related to the virus variant, variations in follow-up duration, immunity profiles, or a combination of these [13]. From a measurement standpoint, the main significance of this finding is that the score can reliably detect differences in HRQoL impact between different cohorts.

### Other data-driven symptom-based post-COVID-19 severity scores

Bahmer et al. previously developed a symptom-based post-COVID-19 severity score [22] to characterise overall syndrome severity using data-driven symptom profiles and demonstrated associations with HRQoL. In their approach, symptom complexes are coded as present or absent, and HRQoL is used as an external validation outcome. In contrast, the present score incorporates graded symptom severity and derives weights directly from their empirical association with HRQoL, thereby quantifying symptom burden in terms of functional health impact rather than relative syndrome severity.

### Applications of the severity score

The continuous nature of the severity score enables dose–response analyses examining associations between biological or clinical markers and post-COVID-19 symptom severity. Instead of relying on simple group comparisons between affected and unaffected individuals, the score facilitates evaluation of graded relations across the entire spectrum of symptom-related HRQoL loss.

Another key intended application of the proposed severity score is the longitudinal assessment of post-COVID-19 burden. By providing a continuous, HRQoL-anchored measure, the score enables the description of heterogeneous recovery and persistence trajectories over time, rather than relying on static case definitions. Such trajectory-based approaches are increasingly recognised as vital for understanding long-term outcomes, identifying subgroups with prolonged impairment, and evaluating the effectiveness of interventions [23,24].

### Strengths and limitations

Several limitations warrant consideration. The severity score is specific to the symptom inventory and severity scaling used in EPILOC and cannot be directly transferred to studies using different instruments without re-estimation of weights. This instrument-specificity, however, is inherent to most symptom-based measures and does not limit the applicability of the underlying HRQoL-anchored approach. Lastly, restricting the score to symptoms not present prior to infection emphasises post-acute burden but may underestimate the overall health impact in individuals with relevant pre-existing conditions. However, our study also has strengths. The score was derived from a large, population-based cohort, enhancing the robustness and generalisability of the findings beyond selected clinical samples. HRQoL was assessed using a well-validated and widely used instrument, ensuring a solid conceptual and methodological foundation for anchoring symptom weights. Moreover, the underlying data included a comprehensive set of post-COVID symptoms assessed using graded severity levels rather than simple presence or absence, allowing for a nuanced representation of symptom burden.

## Conclusions

This study introduces a transparent, HRQoL-anchored symptom severity score that measures graded post-COVID-19 burden beyond simple symptom counts. By empirically linking symptom severity to HRQoL, the score enables a more nuanced assessment of symptom burden and its real-world impact. The method is especially suitable for longitudinal assessment of recovery trajectories and for dose–response analyses in which physiological or biological markers are examined as potential determinants of symptom severity.

## Acknowledgements

We thank key collaborators on this work: Stephan Rusch, Cynthia Stapornwongkul, Jana Kopp, Michaela Schmid, Jennifer Müller, and Patrick Roling.

## Author contributions

Conceptualisation: AN, DR, WVK; Data curation: RSP, LS; Formal Analysis: RSP, LS; Funding acquisition: DR, WVK; Methodology: RSP, CS, LM, AN, DR, WVK; Resources: AN, DR, WVK; Supervision: WVK; Writing – original draft: RSP; Writing – review & editing: LS, AN, CS, LM, SG, UM, JMS, DR, WVK.

## Data availability

The EPILOC consortium has established a Data Access and Use Committee; requests may be sent to.

## Ethics approval

This study was performed in line with the principles of the Declaration of Helsinki. Ethical approval was obtained from the respective ethical review boards of the study centres at Albert-Ludwigs-University Freiburg (Approval No: 21/1484) and Ulm University (Approval No: 337/21).

## Consent to participate

Informed consent was obtained from all individual participants included in the study.

---

**Supplemental Table 1.**
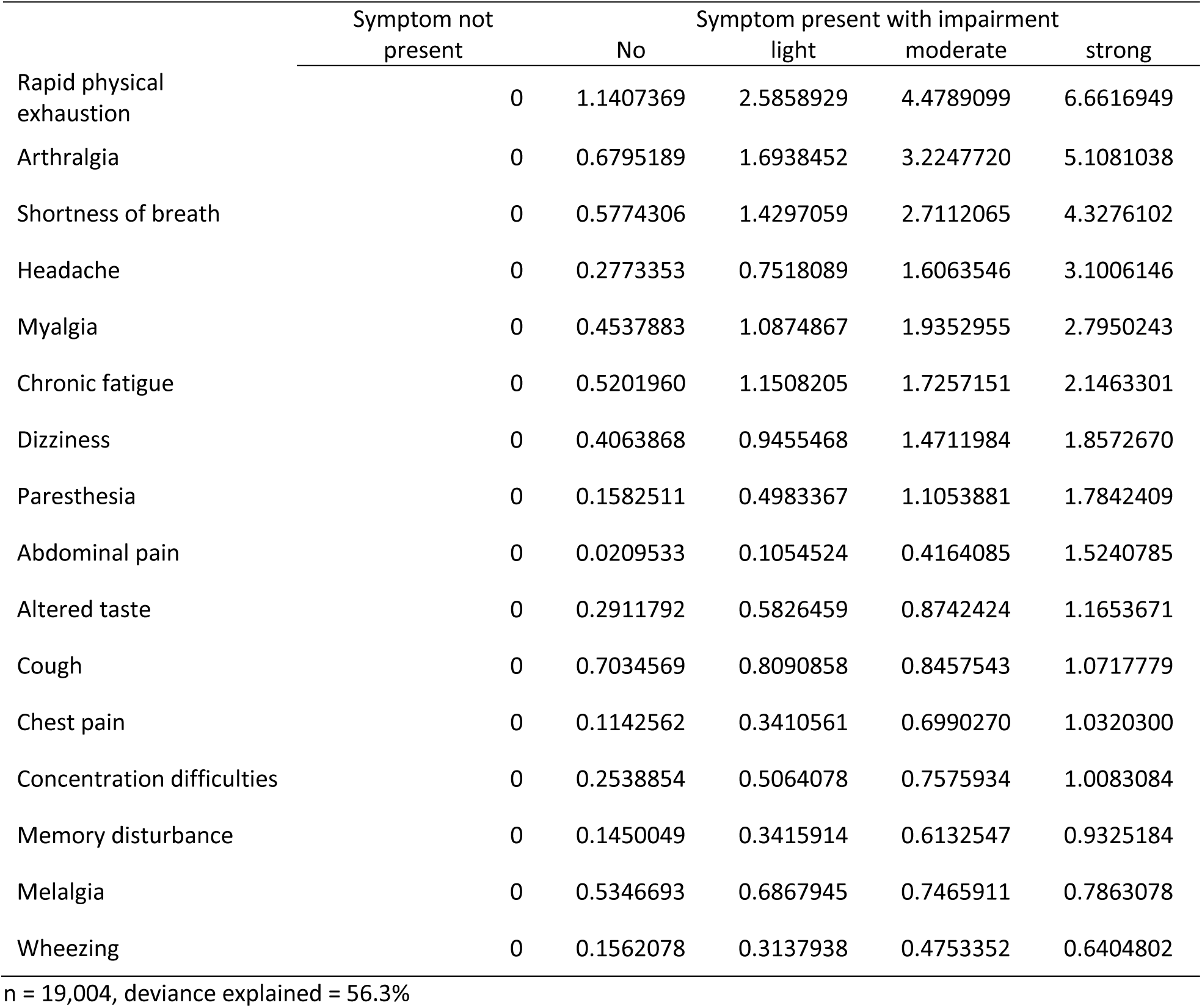
Weights for the physical score – loss in physical HRQoL by grade of symptom severity.

**Supplemental Table 2.**
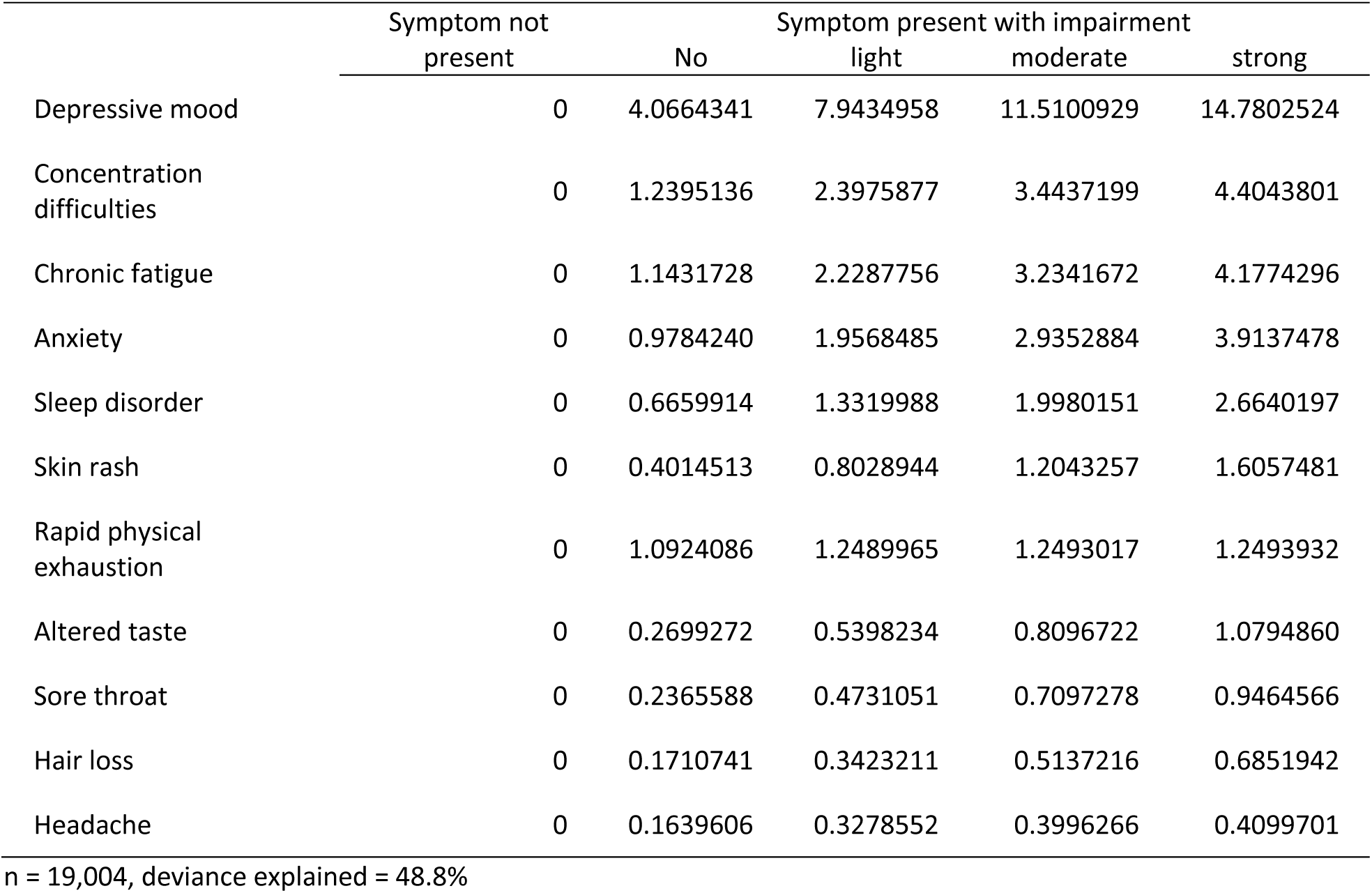
Weights for the mental score – loss in mental HRQoL by grade of symptom severity.

|  | Symptom not present | No | Symptom present with impairment |  |  |
| --- | --- | --- | --- | --- | --- |
|  |  |  | light | moderate | strong |
| Depressive mood | 0 | 4.0664341 | 7.9434958 | 11.5100929 | 14.7802524 |
| Concentration difficulties | 0 | 1.2395136 | 2.3975877 | 3.4437199 | 4.4043801 |
| Chronic fatigue | 0 | 1.1431728 | 2.2287756 | 3.2341672 | 4.1774296 |
| Anxiety | 0 | 0.9784240 | 1.9568485 | 2.9352884 | 3.9137478 |
| Sleep disorder | 0 | 0.6659914 | 1.3319988 | 1.9980151 | 2.6640197 |
| Skin rash | 0 | 0.4014513 | 0.8028944 | 1.2043257 | 1.6057481 |
| Rapid physical exhaustion | 0 | 1.0924086 | 1.2489965 | 1.2493017 | 1.2493932 |
| Altered taste | 0 | 0.2699272 | 0.5398234 | 0.8096722 | 1.0794860 |
| Sore throat | 0 | 0.2365588 | 0.4731051 | 0.7097278 | 0.9464566 |
| Hair loss | 0 | 0.1710741 | 0.3423211 | 0.5137216 | 0.6851942 |
| Headache | 0 | 0.1639606 | 0.3278552 | 0.3996266 | 0.4099701 |

n = 19,004, deviance explained = 48.8%

**Supplemental Figure 1.**
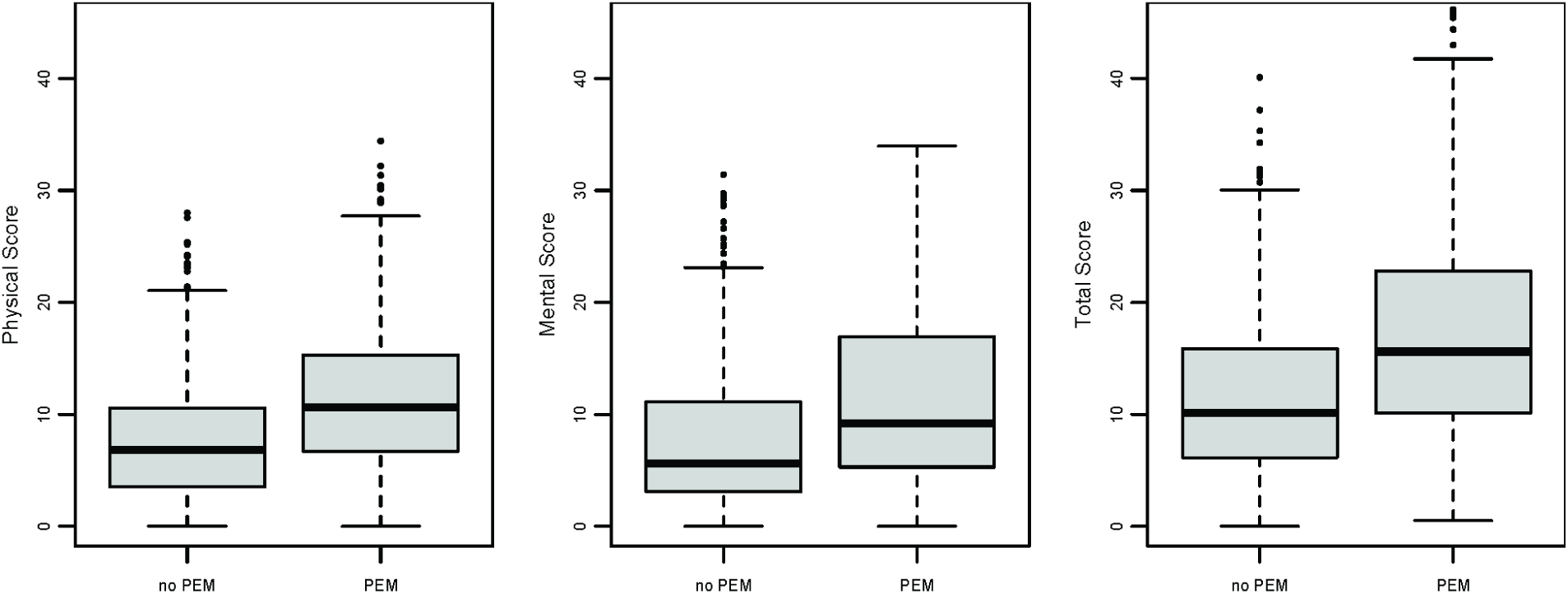
Distribution of symptom severity scores by Post Exertional Malaise (PEM) status in EPILOC Omicron post-covid-19 syndrome cases (n=1242).

